# The Impact of Coronavirus Disease 2019 Pandemic on People with Diabetes in Indonesia: A Cross Sectional National Scale Web-Survey

**DOI:** 10.1101/2020.12.01.20241588

**Authors:** Ida Ayu Kshanti, Marina Epriliawati, Md Ikhsan Mokoagow, Jerry Nasarudin, Nadya Magfira

**Author notes:** Corresponding author: Ida Ayu Kshanti, Emai, Affiliation: Department of Internal Medicine, Fatmawati General Hospital, Indonesia. Jl. RS. Fatmawati Raya No.4, RW.9, West Cilandak., Cilandak, South Jakarta, Jakarta, Indonesia 12430.

## Abstract

**Background:** As the country with the 7th largest number of People with Diabetes (PWD) in the world, the COVID-19 pandemic, and the Large Social Scale Restriction (LSSR) policy taken by the Indonesian government to reduce the number of COVID-19 transmissions is estimated to interfere diabetes management and will increase the incidence of diabetes complications. This study aims to determine the difficulties of diabetes management and its impact on diabetes morbidity during the COVID-19 pandemic in Indonesia.

**Methodology:** This study is a cross-sectional study using a national scale web survey. This research was conducted in Indonesia enrolling 1,124 PWD aged 18 years or older. Diabetes complications are defined as any incidence of hypoglycaemia, or Diabetic Foot Ulcer (DFU), or hospital admission experienced by PWD in Indonesia during the COVID-19 pandemic. The correlation between diabetes management difficulties and diabetes-related complications was measured using a modified cox regression test.

**Results:** Diabetes management difficulties were experienced by 69.8% of PWD in Indonesia. The difficulties include attending diabetes consultation 30.1%, access to diabetes medication 12.4%, checking blood sugar levels 9.5%, controlling diet 23.8%, and performing regular exercise 36.5%. Diabetes-related complications occurred in 24.6% of subjects. Those who had diabetes management difficulties during the COVID-19 pandemic are prone to have diabetes complications by 1.4 times greater (PR: 1.41, 95% CI: 1.09-1.83) than those who did not.

**Conclusion:** The COVID-19 pandemic and LSSR have a substantial impact on diabetes management and indirectly increased diabetes morbidity in Indonesia.

## INTRODUCTION

Coronavirus disease 2019 (COVID-19) caused by the Severe Acute Respiratory Coronavirus Syndrome-2 virus has been declared a public health emergency and global pandemic by WHO on March 11, 2020. [1] COVID-19 has a very diverse clinical spectrum, from asymptomatic to severe symptoms characterized by fever and pneumonia that can be fatal. [2] As of August 11, 2020, COVID-19 has infected 19,936,210 people worldwide with a global Case Fatality Ratio (CFR) of 3.67%. [3] COVID-19 can affect anyone, but subjects with comorbidities such as diabetes have a greater risk of contracting COVID-19 and a higher mortality rate than subjects without diabetes. [4]

From a demographic perspective, Indonesia is the fourth most populous country in the world after China, the United States and India. [5] With a large population, as many as 10.7 million people or 6.2% of the total population are People with Diabetes (PWD), this make Indonesia ranks 7th in the country with the highest number of PWD in the world. [6] If this condition is not handled seriously, Indonesia will remain in the same position in the next 10 years. [6] According to WHO data in 2016, diabetes is one of the main causes of death in Indonesia. [7] In general, diabetes management in Indonesia includes education, dietary management, physical activity, and pharmacological therapy. Comprehensive management in PWD is required to prevent complications.

On March 2, 2020, Indonesia reported the first case of COVID-19. Since the first case discovery until August 11, 2020, 130,718 people in Indonesia have been diagnosed with COVID-19 and 4.5% of these cases have died (5,880 people). [3] As an effort to deal with pandemics and public health emergencies, Indonesia has imposed a regional quarantine in the form of large-scale social restrictions (LSSR) on affected cities and provinces. The Indonesian Society of Endocrinology (ISE/ PERKENI) has issued recommendations for PWD to stay at home and maintain physical distance to reduce exposure to viral carriers. [8] However, this policy poses an unfavourable impact on the management of diabetes. Face-to-face consultations are largely avoided. The fear of contracting COVID 19 are present both in patients and some health care providers. LSSR have rendered patients physically less active or unable to exercise. Also, maintaining the required dietary becomes more difficult. In a worst-case scenario oral anti-diabetic drugs and/or insulin were more difficult to obtain. [9] The impact of this policy may bring about more severe impact on PWD than the COVID 19 disease *per se*.

This study assesses the wider impact of COVID-19 pandemic beyond the effects of the disease itself. We assessed the impact of social restriction policies and the enactment of COVID-19 as a public health emergency, particularly its impact on PWD. Furthermore, this research may provide basis for decision making by the government to optimize diabetes management, especially during the COVID-19 pandemic in Indonesia. The purpose of this study was to determine the difficulties faced by PWD during the COVID-19 pandemic in Indonesia and their impact on morbidity. This study also assesses the solutions taken by PWD in response to difficulties on diabetes management during the COVID-19 pandemic.

## METHODS

### 1. Study Design and Sample

This study is a cross-sectional study using a national scale web-survey. Flyers containing invitations to become research respondent were given to the professional organization (ISE, Indonesian Medical Association, Indonesia Society of Internal Medicine, Indonesia Diabetes Association and Indonesia Diabetes Educators Association) which would then be given to PWD. The survey was conducted for two weeks during the period of 21 July 2020-4 August 2020. This study involved all PWD in Indonesia who lived in Indonesia during March - July 2020 and were not being hospitalized when filling out the questionnaire. To find out the relationship between difficulties during the COVID-19 pandemic and the incidence of complications during the COVID-19 pandemic with a study power of 95% and a 95% confidence interval, a sample of 970 diabetes mellitus patients was needed.

### 2. Research Variables and Outcomes

Difficulties during the pandemic period were defined as any kind of difficulties that were subjectively endured/experienced by the PWD (difficulties in attending health consultation, or checking blood sugar levels, or obtaining diabetes drugs, or maintaining diet control, or performing exercise). Complications were defined as any experiences of hypoglycaemia or Diabetes Foot Ulcers (DFU) or hospitalisation during the COVID-19 pandemic in Indonesia.

### 3. Data Sources

This study uses primary data in the form of a questionnaire containing gender, age, occupation, income, health insurance, transportation, residence, and distance to the health facility. Working condition were classified into work from home and work from the office. The distance to the health facility is a subjective measure of distance from home to the control place according to the subject. This study also measured diabetes profile which included duration of diagnosis, diabetes treatment, diabetes control facility, and numbers of medical consultation during the pandemic.

### 4. Statistical methods

The relationship between difficulties and the incidence of complications during the COVID-19 pandemic was measured by assessing the prevalence ratio (PR) and 95% confidence interval (CI) using a modified cox-regression test.

## RESULTS

### 1. Characteristics of Subjects

This study included 1,124 PWD from 34 provinces in Indonesia (Figure 1). The characteristics of the subjects in this study can be seen in Table 1. The proportion of male includes 54.89% of the total respondents, almost 60% of the respondents are in productive age, less than a quarter of the respondents have an income below the National Minimum Wage (NMW), and almost 80% of the total respondents receive treatment using National Health Insurance (NHI)/ *Badan Penyelenggara Jaminan Sosial* (BPJS). In this study, the majority of patients were married and lived with their families. As many as 35% of patients felt that they live far away from the health facility where they regularly attended for diabetes treatment. Also, the majority of patients went to the facility using private vehicles.

**Table 1.** Demographic Characteristics of the Subjects

| Variable | Frequency (n) | Presentation (%) |
| --- | --- | --- |
| **Sex** |  |  |
| Male | 507 | 54.89 |
| Female | 617 | 45.11 |
| **Age** |  |  |
| 18-40 years | 144 | 12.81 |
| > 40-60 years | 514 | 45.73 |
| > 60 years | 466 | 41.46 |
| **Occupation** |  |  |
| Does Not Work | 604 | 53.74 |
| Work from Home | 259 | 49.81 |
| Work from Office | 261 | 50.19 |
| **Income** |  |  |
| < Rp. 2.000.000 | 238 | 21.17 |
| Rp. 2.000.000-Rp.5.000.000 | 464 | 41.28 |
| Rp. 5.000.000-Rp.10.000.000 | 208 | 18.51 |
| > Rp. 10.000.000 | 214 | 19.04 |
| **Health Insurance** |  |  |
| National Health Insurance | 880 | 78.30 |
| Other Insurance | 78 | 6.94 |
| Pay with Own Money | 64 | 5.69 |
| **Marriage** |  |  |
| Married | 978 | 87.01 |
| Single | 43 | 3.83 |
| Ever been married | 103 | 9.16 |
| **Transportation to Health Facility** |  |  |
| Private vehicle | 861 | 76.60 |
| By foot | 16 | 1.42 |
| Online Transportation | 106 | 9.43 |
| Public transportation | 141 | 12.54 |
| **Residence** |  |  |
| Live with family | 936 | 83.27 |
| Live alone | 183 | 16.28 |
| Live in a boarding house/ institution | 5 | 0.44 |
| **Distance to Health Facility** |  |  |
| Close | 731 | 65.04 |
| Far | 393 | 34.96 |

\*Rp: Rupiah (IDR)

**Figure 1.**
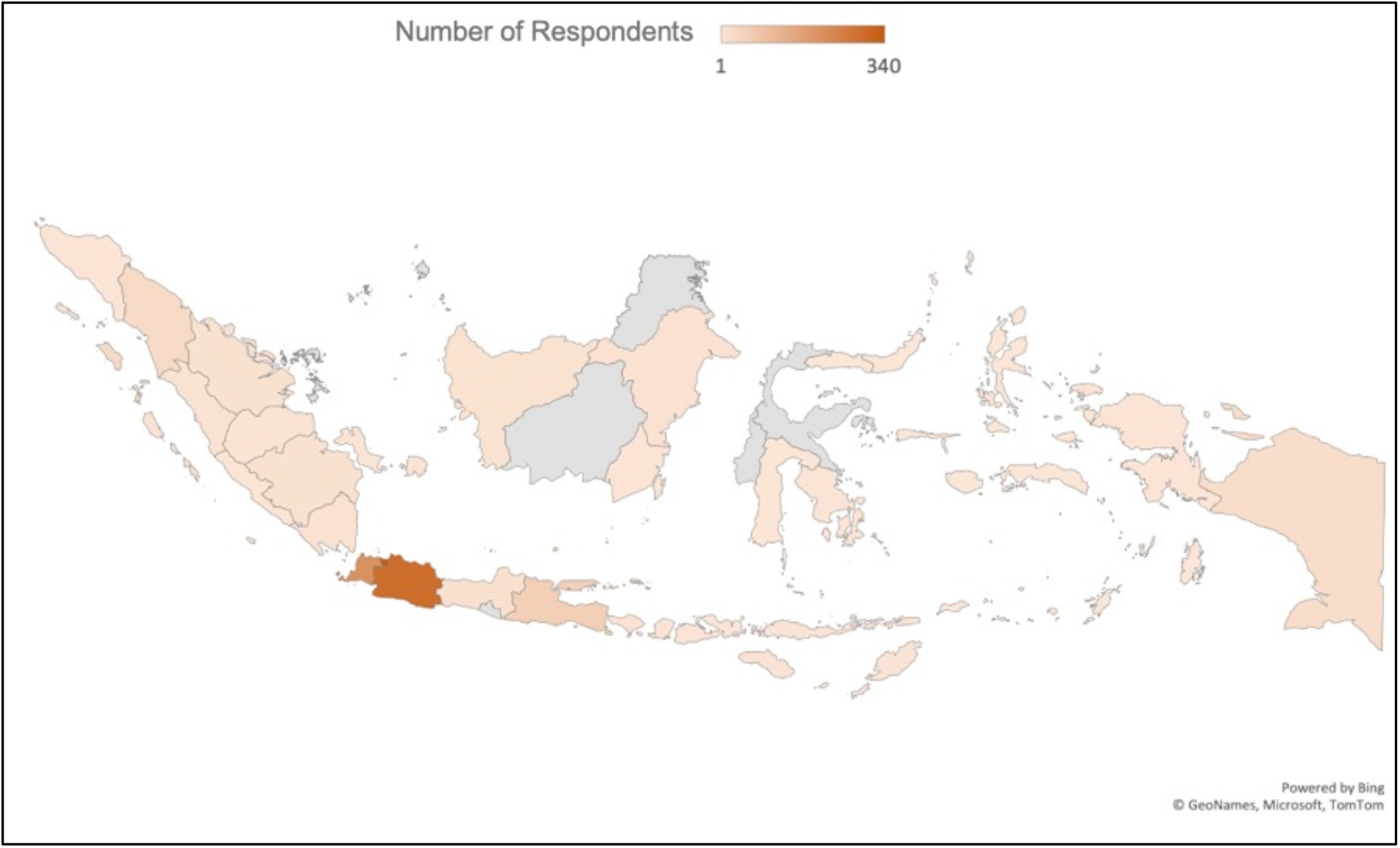
Distribution of Subjects According to Regions in Indonesia.

The diabetes profile of the subjects can be seen in Table 2. The majority of respondents had been diagnosed with diabetes for more than three years and visited a hospital for diabetes treatment. As many as 74.02% of respondents received oral anti-diabetic drug therapy and as many as 40.3% of patients received insulin therapy. In this study, 77.85% of the total number of respondents usually had, at least, one consultation in three months before the COVID-19 pandemic.

**Table 2.** Diabetes Profile

| Variable | Frequency (n) | Presentation (%) |
| --- | --- | --- |
| **Diabetes Duration** |  |  |
| < 3 years | 253 | 22.51 |
| ≥ 3-5 years | 257 | 22.86 |
| ≥ 5-10 years | 233 | 20.73 |
| ≥ 10 years | 381 | 33.90 |
| **Diabetes Medication** |  |  |
| Do not know | 21 | 1.87 |
| No pharmacological treatment | 56 | 4.98 |
| OAD | 594 | 52.85 |
| Insulin | 215 | 19.13 |
| OAD and Insulin | 238 | 21.17 |
| **Health Facility for Diabetes Treatment** |  |  |
| None | 87 | 7.74 |
| Hospital | 763 | 67.88 |
| Public health centre | 64 | 5.69 |
| Private clinic | 122 | 10.85 |
| Private practice | 86 | 7.65 |
| Midwife / nurse | 2 | 0.18 |
| **Number of regular consultations before the pandemic** |  |  |
| More than once in a month | 30 | 2.67 |
| Once in a month | 657 | 58.45 |
| Once in 2 months | 69 | 6.14 |
| Once in 3 months | 119 | 10.59 |
| Once in 6 months | 93 | 8.27 |
| Once in a year | 71 | 6.32 |
| Never | 85 | 7.56 |

OAD: Oral Anti Diabetics

### 2. Impact of the COVID-19 Pandemic on Diabetes Morbidity

In this study, 24.56% of the total respondents admitted that they had diabetes complications during pandemic (hypoglycaemia 12.90%, DFU 7.38%, and hospital admission 6.76%). About 70% of PWD experienced difficulties during the COVID-19 pandemic, which include attending diabetes consultation (30.07%), access to diabetes medication (12.37%), checking blood sugar levels (9.52%), controlling diet (23.75%) and performing regular exercise (36.48%). Multivariate analysis results show an increased incidence of diabetes complications by 1.41 times during the COVID 19 pandemic (95% CI: 1.09-1.83) (Table 3). Other factors that play a role on increasing number of incidences were found in PWD under 60 years of age (PR: 1.44, 95% CI: 1.12-1.85) and those receiving insulin therapy (PR: 2.23, 95% CI: 1.75-2.85).

**Table 3.** Relationship between Difficulties During the COVID-19 Pandemic and Diabetes Complications in People with Diabetes in Indonesia

| Variable | Complications n (%) |  | PR Crude<br>(95% CI) | PR Adjusted (95%<br>CI) |
| --- | --- | --- | --- | --- |
|  | Present<br>276 (24.56) | Not Present<br>848 (75.44) |  |  |
| **Difficulties in diabetes management** |  |  |  |  |
| Present | 194 (70.29) | 485 (57.19) | 1.55 (1.23-1.95) * | 1.41 (1.09-1.83) |
| Not Present | 82 (29.71) | 363 (42.81) | 1.00 | 1.00 |
| **Sex** |  |  |  |  |
| Male | 124 (44.93) | 383 (45.17) | 0.99 (0.81-1.22) |  |
| Female | 152 (55.07) | 465 (54.83) | 1.00 |  |
| **Age** |  |  |  |  |
| < 60 years | 186 (67.39) | 472 (55.66) | 1.46 (1.17-1.83) * | 1.44 (1.12-1.85) |
| ≥ 60 years | 90 (32.61) | 376 (44.34) | 1.00 | 1.00 |
| **Occupation** |  |  |  |  |
| Work from Office | 54 (41.86) | 207 (52.94) | 0.71 (0.53-0.97) * |  |
| Work from Home/<br>Unemployed/Retired | 75 (58.14) | 184 (47.06) | 1.00 |  |
| **Income** |  |  |  |  |
| Below NMW | 73 (26.45) | 165 (19.46) | 1.34 (1.07-1.68) * |  |
| NMW/ above NMW | 203 (73.55) | 683 (80.54) | 1.00 |  |
| **Health Insurance** |  |  |  |  |
| NHI | 38 (13.77) | 104 (12.26) | 1.10 (0.82-1.48) |  |
| Other | 238 (86.23) | 744 (87.74) | 1.00 |  |
| **Residence** |  |  |  |  |
| Live alone | 46 (16.67) | 137 (16.16) | 1.03 (0.78-1.35) |  |
| Live with family/ other | 230 (83.33) | 711 (83.84) | 1.00 |  |
| **Distance to Health Facility** |  |  |  |  |
| Far | 109 (39.49) | 284 (33.49) | 1.21 (0.99-1.50) |  |
| Close | 167 (60.51) | 564 (66.51) | 1.00 |  |
| **Diabetes duration** |  |  |  |  |
| ≥ 5 years | 163 (59.06) | 451 (53.18) | 1.20 (0.97-1.48) |  |
| < 5 years | 113 (40.94) | 397 (46.82) | 1.00 |  |
| **Diabetes medications** |  |  |  |  |
| Insulin | 168 (60.87) | 285 (33.61) | 2.30 (1.87-2.84) * | 2.24 (1.75-2.85) |
| Others | 108 (39.13) | 563 (66.39) | 1.00 | 1.00 |
| **Health Facility for Diabetes Treatment** |  |  |  |  |
| Hospital | 188 (68.12) | 575 (67.81) | 1.01 (0.81-1.26) |  |
| Others | 88 (31.88) | 273 (32.19) | 1.00 |  |
| **Number of regular consultations before the pandemic** |  |  |  |  |
| ≥ Once in 3 months | 59 (21.38) | 190 (22.41) | 0.96 (0.74-1.23) |  |
| < Once in 3 months | 217 (78.62) | 658 (77.59) | 1.00 |  |

### 3. How People with Diabetes in Indonesia Cope with Difficulties of Diabetes Management During Pandemic

The study shows that, the majority of PWD did not take any action in maintaining/resolving their diabetes health-related problems during the COVID pandemic in Indonesia. These include as many as 50.59% of respondents who had difficulty in attending diabetes consultation, 60.75% who had difficulty in checking blood sugar levels, and 30.94% who had difficulty in obtaining anti-diabetes drugs or insulin. (Table 4).

**Table 4.** How People with Diabetes in Indonesia Cope with Difficulties of Diabetes Management During Pandemic

| Variables | Frequency (n) | Presentation (%) |
| --- | --- | --- |
| **Difficulties in attending diabetes consultation** | 338 | 30.7 |
| Disregard the condition/indifference | 171 | 50.59 |
| Chat with health providers via non health applications | 62 | 18.34 |
| Call health providers for a consultation | 32 | 9.47 |
| Consultation with health providers via health applications/ internet | 73 | 21.60 |
| **Difficulties in checking blood sugar levels** | 107 | 9.52 |
| Left alone (do not check blood sugar levels) | 65 | 60.75 |
| Buy a glucometer | 35 | 32.71 |
| Borrow glucometer from friends / relatives | 7 | 6.54 |
| **Difficulties in obtaining diabetes medications** | 139 | 12.37 |
| Left alone (do not take medicine) | 43 | 30.94 |
| Buy medicine at pharmacies / online / internet applications | 78 | 56.12 |
| Ask friends / family to buy medicine | 11 | 7.91 |
| Change medication without consultation | 5 | 3.60 |
| Consult a doctor / educator / nurse via application / internet to get a prescription | 2 | 1.44 |

## DISCUSSION

This study shows that nearly 70% of PWD in Indonesia experienced difficulties in managing diabetes during the COVID-19 pandemic. As many as 24.6% of these subjects experienced complications related to their diabetes including DFU (7.4%), hypoglycaemia (12.9%), and hospitalisation (6.8%). In this study, subjects who experienced difficulties during the COVID-19 pandemic had an increased risk to develop diabetes complications by 1.4 times more often than those who did not during the COVID-19 pandemic.

In dealing with the difficulties afflicted by PWD during the COVID-19 pandemic in Indonesia, only less than half of the respondents took the initiative to utilize available online resources/information or to contact a doctor for consulting their health conditions. However, more than half of respondents answered that they did nothing and just let their condition deteriorate. Moreover, more than a third of PWD discontinue their medication during the COVID-19 pandemic.

This study has some limitation including reporting of complication during pandemic limited to DFU, hospital admission, or hypoglycaemia. The Questionnaire was answered subjectively by respondents. Hence, the observations may have had some biases. Furthermore, the condition of unawareness towards DFU and undetected/undocumented low or subclinical blood sugar levels, which were not assessed objectively, could result in an overall lower number of complications than what actually occurred.

This study shows that the COVID-19 pandemic in Indonesia has had a substantial impact on the management of diabetes. At least one in four PWD experienced complications during the COVID-19 pandemic in Indonesia. In this study, the prevalence of DFU in diabetics in Indonesia was found in 7.4% of the total respondents. Until now, there is no national data on the prevalence of diabetes complications in Indonesia. However, a single-centre study showed the prevalence of DFU incidence in the population treated at a tertiary hospital in Indonesia was 16.7% with an amputation rate of 10.7%. [10] Meanwhile, the worldwide prevalence rate for DFU is 6.3% (95% CI: 5.4% −7.3%). [11] Given the limitations of this study, the total prevalence of DFU in diabetes in Indonesia is likely higher than what was found in this study. However, it can be concluded that when compared with the prevalence rates for DFUs worldwide, the prevalence of DFU in Indonesia during the pandemic period was higher.

In this study, the incidence of hypoglycaemia was reported at 12.9% in all subjects. Worldwide, the incidence of hypoglycaemia is 45% for mild-moderate hypoglycaemia and 6% for severe hypoglycaemia. [12] In a cohort study held in Indonesia, the incidence of hypoglycaemia was found in 99.4% of subjects with Type 2 Diabetes Mellitus (T2DM) (59% severe) and 100% in subjects with Type 1 Diabetes Mellitus (T1DM) (76% severe). [13] Interestingly, as many as 62.17% of patients with T1DM and 82.4% of T2DM patients in Indonesia were unaware of their hypoglycaemic conditions. [13] Apart from the fact that the hypoglycaemia events in this study is subjective, the lack awareness of hypoglycaemia in PWD in Indonesia could explain why the incidence of hypoglycaemia in our study is low.

When viewed from the perspective of hospital admission, the incidence of hospitalisation during the pandemic among PWD in this study was 6.76%. When compared to COVID-19 cases, 18% of patients experienced severe clinical manifestations and required hospitalization while severe cases occurred in 44% of COVID-19 patients with diabetes comorbidity.[4] Although the hospitalisation rate for PWD in this study was quite low, actual numbers may have been higher due to the limited timeframe during which the data were acquired (4 months since pandemic began). In addition, lower hospitalisation rate may have also arisen from selection bias of the study.

During the COVID-19 pandemic, the majority of PWD experienced difficulties in managing their disease. These various difficulties increased the incidence of diabetes complications for almost 1.5 times. At present, although the number of cases and deaths due to COVID-19 continues to increase, the death and cases of diabetes in Indonesia are still much higher than COVID-19. [3,7] Putting aside comprehensive management of chronic diseases and mainly focusing on COVID-19 may potentially be harmful. For this reason, rapid and sustainable changes in the health sector to reduce the impact of COVID-19 pandemic on general health specifically in chronic disease is needed. [14]

Virtual consultation that eliminates the need for physical meetings is a solution that has been implemented by many countries in the world, including Indonesia. [15] In Indonesia, ISE has provided recommendations to avoid direct consultation and suggested remote consultation for diabetic patients when applicable. [8] However, only less than half of the PWD in Indonesia used it, while the remaining subjects remained idle, allowing their condition to deteriorate. In contrast, since the COVID-19 pandemic, doctors in People’s Republic of China have shifted their services to telemedicine, and the number of patients being served are more than 100 people per day per doctor, which exceeds the number that could be handled before the pandemic era. [15] The United States, Italy, and South Africa are also implementing the same approach, and even audio-only telemedicine service is provided as one solution although its effectiveness is still under study in the United States. [16]

The concept of telemedicine is not new, even though the quality of virtual services will not be the same as direct meetings and physical examinations, changes due to the COVID-19 pandemic must be made because we still don’t know when this pandemic will end. [14, 19] To create wide coverage of telemedicine services in Indonesia, various barriers are identified; as an archipelago country, health services in Indonesia are not evenly distributed.[17] The health expenditure is less than 5% and the universal health coverage does not yet cover the entire population. [17] Acknowledging infrastructure condition of information and technology, 98% of fibre optic backbone is available on the Island of Java, but it is not yet in the island of Maluku-Papua. Only 66.2% of households in Indonesia have access to the internet and only 39.90% of individuals use the internet. However, as an opportunity, 121 out of 100 residents are cellular telephone users, this means that 21 residents have more than one cellular contact. [18]

Although many are unprepared in facing these changes, the collaboration between the government in providing infrastructure and health service providers including clinicians as the vanguard is necessary. The use of audio-only telemedicine may be applicable in Indonesia, especially in provinces with inadequate internet access. Further research is certainly needed to assess the effectiveness and implementation of telemedicine in health services during the COVID-19 pandemic in Indonesia and how it will be sustainable in the future.

However, beyond the readiness of the government and health service providers in developing telemedicine, it is also important to improve public health literacy. Improving health literacy requires collaborative efforts, involving various stakeholders. A person with poor health literacy has a poor health status as well. [20] Therefore, good health literacy is needed by PWD to manage their health and make the right health decisions to achieve favourable disease outcomes. [21, 22] Health literacy in diabetes can be improved by increasing knowledge on self-efficacy and self-care in diabetes management. [23, 24] Utilizing technology to facilitate education on various aspects of diabetes management especially during the pandemic may offer solution to improve health literacy in the society. [25]

## CONCLUSION

The results of this study may be generalized to all diabetes patients in Indonesia and in other countries whose demographics, economies, and geographies are similar to Indonesia. This research can ultimately provide recommendations to various stakeholders including clinicians and PWD in the form of (1) the COVID-19 pandemic has substantial impact on diabetes management and increases diabetes complications, (2) the use of telemedicine in diabetes management needs to be strengthened and may offer a solution to overcome difficulties experienced by PWD during the COVID-19 pandemic, (3) cooperation between the government and health service providers in providing telemedicine services for PWD is necessary, (4) health organizations and government need to collaborate to formulate standards or guidelines for diabetes services during a pandemic and, (5) improving health literacy with technology-based health promotion may provide a solution to reduce the incidence of diabetes-related complications during a pandemic.

## Data Availability

The data that support the findings of this study are available from the corresponding author, IAK, upon reasonable request

## DECLARATIONS

### Ethical approval

This study was approved by the research ethics committee of Fatmawati General Hospital 11/KPP/VII/2020. Informed consent was obtained from the respondent via electronic approval.

### Consent for publication

Not applicable

### Availability of data and materials

The original data from this study and the analysed results will be available from the corresponding author upon reasonable request

## LIST OF ABBREVIATIONS

COVID 19: Coronavirus disease 2019
CI: Confidence Interval
DFU: diabetic foot ulcer
ISE: Indonesian Society of Endocrinology
LSSR: large social scale Restriction
NHI: National Health Insurance
NMW: National Minimum Wage
OAD: Oral Anti Diabetics
PR: prevalence ratio
PWD: People with Diabetes
RP: Rupiah

## Acknowledgements

Not applicable

## Author contributions

IAK, contributed to development of study concept and designs. IAK, ME, MI, JN managed the overall project and were responsible for the questionnaire survey of people in Indonesia. NM contributed to analysis and interpretation of data. MI, NM contribute to drafting of the first draft. IAK, ME, JN finalized the manuscript on the basis of comments from other authors. All authors read and approved the final manuscript.

## References

1. World Health Organization (WHO): Coronavirus disease 2019 (COVID-19) Situation Report-67. 2020.

2. Liu Y, Yan L-M, Wan L, Xiang T-X, Le A, Liu J-M, Peiris M, Poon LLM, Zhang W: Viral dynamics in mild and severe cases of COVID-19. The Lancet Infectious Diseases 2020, 20(6):656–657.

3. World Health Organization (WHO): Coronavirus Disease (COVID-19) Situation Report-204. 2020.

4. Sanyaolu A, Okorie C, Marinkovic A, Patidar R, Younis K, Desai P, Hosein Z, Padda I, Mangat J, Altaf M: Comorbidity and its Impact on Patients with COVID-19. SN Comprehensive Clinical Medicine 2020, 2(8):1069–1076.

5. Elias S, Noone C: The Growth and Development of The Indonesian Economy. Bulletin 2011, December Quarter 2011.

6. International Diabetes Federation (IDF): IDF Diabetes Atlas Ninth Edition 2019. International Diabetes Federation 2019.

7. World Health Organization (WHO): Diabetes Country Profile: Indonesia. World Health Organization 2016.

8. Perkumpulan Endokrinologi Indonesia (PERKENI): Pernyataan Resmi dan Rekomendasi Pengangan Diabetes Mellitus di era Pandemi COVID-19. PERKENI 2020, IV(239).

9. Ghosh A, Gupta R, Misra A: Telemedicine for diabetes care in India during COVID19 pandemic and national lockdown period: Guidelines for physicians. Diabetes Metab Syndr 2020, 14(4):273–276.

10. Pemayun TGD, Naibaho RM: Clinical profile and outcome of diabetic foot ulcer, a view from tertiary care hospital in Semarang, Indonesia. Diabet Foot Ankle 2017, 8(1):1312974.

11. Zhang P, Lu J, Jing Y, Tang S, Zhu D, Bi Y: Global epidemiology of diabetic foot ulceration: a systematic review and meta-analysis (dagger). Ann Med 2017, 49(2):106–116.

12. Edridge CL, Dunkley AJ, Bodicoat DH, Rose TC, Gray LJ, Davies MJ, Khunti K: Prevalence and Incidence of Hypoglycaemia in 532,542 People with Type 2 Diabetes on Oral Therapies and Insulin: A Systematic Review and Meta-Analysis of Population Based Studies. PLoS One 2015, 10(6):e0126427.

13. Rudijanto A, Saraswati MR, Yunir E, Kumala P, Puteri HHS, Mandang VVV: Indonesia Cohort of IO HAT Study to Evaluate Diabetes Management, Control, and Complications in Retrospective and Prospective Periods Among Insulin-Treated Patients with Type 1 and Type 2 Diabetes. Acta Medica Indonesiana 2018, 50(1).

14. Alromaihi D, Alamuddin N, George S: Sustainable diabetes care services during COVID-19 pandemic. Diabetes Res Clin Pract 2020, 166:108298.

15. Webster P: Virtual health care in the era of COVID-19. The Lancet 2020, 395(10231):1180–1181.

16. Center for Medicare and Medicaid Service: Physicians and Other Clinicans: CMS Flexibilities to Fight COVID-19. Center for Medicare and Medicaid Service 2020.

17. Indonesia MoHRo: Indonesia Health Profile 2018. Ministry of Health Republic of Indonesia 2019.

18. Badan Pusat Statistik: ICT Development Index 2018. BPS-Statistic Indonesia 2019.

19. Klonoff DC: Telemedicine for Diabetes After the COVID-19 Pandemic: We Can’t Put the Toothpaste Back in the Tube or Turn Back the Clock. J Diabetes Sci Technol 2020, 14(4):741–742.

20. Al Sayah F, Majumdar SR, Williams B, Robertson S, Johnson JA: Health literacy and health outcomes in diabetes: a systematic review. J Gen Intern Med 2013, 28(3):444–452.

21. Abdullah A, Liew SM, Salim H, Ng CJ, Chinna K: Prevalence of limited health literacy among patients with type 2 diabetes mellitus: A systematic review. PLoS One 2019, 14(5):e0216402.

22. Olesen K, AL FR, Joensen L, Ridderstrale M, Kayser L, Maindal HT, Osborne RH, Skinner T, Willaing I: Higher health literacy is associated with better glycemic control in adults with type 1 diabetes: a cohort study among 1399 Danes. BMJ Open Diabetes Res Care 2017, 5(1):e000437.

23. Cavanaugh KL: Health literacy in diabetes care: explanation, evidence and equipment. Diabetes Manag (Lond) 2011, 1(2):191–199.

24. Landry KE: Using eHealth to improve health literacy among the patient population. Creat Nurs 2015, 21(1):53–57.

25. Evers KE: eHealth promotion: the use of the Internet for health promotion. Am J Health Promot 2006, 20(4):suppl 1-7, iii.

